# Design and feasibility of an Alzheimer’s disease blood test study in a diverse community-based population

**DOI:** 10.1101/2023.01.31.23285249

**Authors:** Melody Li, Yan Li, Suzanne E. Schindler, Daniel Yen, Siobhan Sutcliffe, Ganesh M. Babulal, Tammie L.S. Benzinger, Eric J. Lenze, Randall J. Bateman

**Affiliations:** Department of Neurology, Washington University School of Medicine, St. Louis, MO, 63110, USA; The Tracy Family Stable Isotope Labeling Quantitation Center for Neurodegenerative Biology, Washington University School of Medicine, St. Louis, MO, 63110, USA; Hope Center for Neurological Disorders, Washington University School of Medicine, St. Louis, MO, 63110, USA; Knight Alzheimer Disease Research Center, Washington University School of Medicine, St. Louis, MO, 63110, USA; Department of Surgery – Public Health Sciences, Washington University School of Medicine, St. Louis, MO, 63110, USA; Mallinckrodt Institute of Radiology, Washington University School of Medicine, St. Louis, MO, 63110, USA; Department of Psychiatry, Washington University School of Medicine, St. Louis, MO, 63110, USA

**Keywords:** Alzheimer’s disease, dementia, cognitive impairment, blood-based biomarkers, blood test, amyloid PET, clinical trial enrollment, recruitment

## Abstract

**INTRODUCTION:** Alzheimer’s disease (AD) blood tests are likely to become increasingly important in clinical practice, but need to be evaluated in diverse groups before use in the general population.

**METHODS:** This study enrolled a community-based sample of older adults in the Saint Louis, Missouri, USA area. Participants completed a blood draw, AD8® dementia screening interview, Montreal Cognitive Assessment (MoCA), and survey about their perceptions of the blood test. A subset of participants completed additional blood collection, amyloid PET, MRI, and Clinical Dementia Rating® (CDR).

**RESULTS:** Of the 859 participants enrolled in this ongoing study, 20.6% self-identified as Black or African American. The AD8 and MoCA correlated moderately with the CDR. The blood test was well-accepted by the cohort, but perceived more positively by White and highly educated individuals.

**DISCUSSION:** Studying an AD blood test in a diverse population is feasible, and may accelerate accurate diagnosis and implementation of effective treatments.

## 1. Introduction

Alzheimer’s disease (AD) is the most common cause of dementia, and associated pathological changes begin a decade or more before symptom onset [1–3]. Measures of amyloid-β (Aβ) and phosphorylated tau (p-tau) species in cerebrospinal fluid (CSF), as well as positron emission tomography (PET) scans using radiotracers that bind to aggregated amyloid and tau, are highly accurate biomarkers of AD pathology [3–6]. However, lumbar punctures may be perceived as invasive, particularly among underrepresented groups in AD research [7,8], while PET scans are expensive, involve exposure to low levels of radiation, and only available at specialized medical centers. Recent advances in the development of AD blood tests provide a simple and cost-effective way to detect AD pathology and have the potential to increase access of minoritized groups to AD biomarker testing [9]. Plasma Aβ42/40 measurement by mass spectrometry has been validated in archived samples from longitudinal observational research studies of AD [10–15], and is currently available for use in clinical diagnosis. Biomarkers that accurately detect AD brain pathology, including in cognitively normal individuals, may accelerate development of effective treatments and enable more accurate clinical diagnoses.

Although longitudinal AD research cohorts are deeply phenotyped with rich clinical, cognitive, and biomarker data, these cohorts are typically not representative of the broader population. AD research studies often lack significant ethnoracial, socioeconomic, and comorbid diversity [16,17]. Minoritized groups have a higher risk of AD dementia [18–20], making it especially problematic that they are underrepresented in AD research [21]. Several studies have found racial differences in AD biomarkers, including lower average CSF total tau and p-tau181 concentrations in Black or African American compared to non-Hispanic White participants [22–25]. Although AD blood tests may perform differently across racial groups in predicting AD pathology [25], this is difficult to clearly establish because so few individuals from minoritized groups have samples available for analysis [26,27]. Certain health conditions such as chronic kidney disease may also affect the results of AD blood tests [28,29]. AD blood tests are likely to play an increasingly important role in clinical practice, especially with implementation of anti-amyloid drugs approved by the Food and Drug Administration [30,31]. Therefore, it is imperative to evaluate whether AD blood tests perform consistently and accurately in diverse, community-based samples to determine whether they can be used in the general population for clinical trial screening and clinical diagnosis.

In this context, we launched the Study to Evaluate Amyloid in Blood and Imaging Related to Dementia (SEABIRD) in 2019, an observational study to collect blood samples from 1,120 participants mirroring the demographics of the greater Saint Louis metropolitan area. Here, we report on the first 859 participants enrolled in this ongoing study. We examine the demographics, medical conditions, and cognitive characteristics of this community-based population. We demonstrate the feasibility and acceptability of a community-based blood collection study despite the COVID-19 pandemic, and explore differences in attitudes toward AD blood tests among participant groups.

## 2. Methods

### 2.1. Study Population

SEABIRD is a cross-sectional observational study of community-dwelling older adults in the greater metropolitan area of Saint Louis, Missouri, USA. The Washington University institutional review board approved the study, and all participants provided written informed consent.

Eligible participants were aged 60 years or older and either cognitively normal or exhibited mild cognitive impairment as defined by an abnormal score on the AD8® dementia screening interview and/or the Montreal Cognitive Assessment (MoCA) [32,33]. Those who reported inability to perform one or more activities of daily living (eating, bathing, dressing, ambulating, toileting) due to advanced cognitive impairment were excluded. Additional exclusion criteria included active infectious disease, presence of a bleeding disorder, or use of an experimental drug for AD.

Participants were recruited from a variety of sources, including newspaper advertisements and local news features, word of mouth (snowball sampling), electronic medical record (EMR) reviews, social media advertisements, participant registries and collaborating studies, community outreach, and distribution of study flyers to local organizations. Recruitment was guided by the study’s Data and Safety Monitoring Committee (DSMC), which reviewed enrollment progress and advised on recruitment strategies to reach underrepresented groups. Strategies included advertisements tailored for specific groups, prioritization of minoritized group enrollment, a waitlist for over-represented groups, and protocol changes based on participant feedback (e.g., increased remuneration for study visits).

Planned enrollment numbers for demographic characteristics were estimated from the 2020 American Community Survey 5-Year Estimates tables for the population 60 or 65 years and over in the St. Louis, MO-IL Metro Area [34–36]. Planned enrollment numbers for medical conditions, apolipoprotein E (*APOE*) ε4 prevalence, and cognitive impairment were determined from a 2013 Missouri Department of Health and Senior Services report on the prevalence of chronic diseases in Missouri older adults aged 65+, the 2022 Alzheimer’s Disease Facts and Figures Alzheimer’s Association Report, and results from nationally representative studies, respectively [37–40].

### 2.2 Study Procedures

Participants were screened for study eligibility over the phone. During the initial phone call, a research coordinator completed a dementia screening interview (AD8) with an individual who knew the participant well (informant). If no informant was available, the AD8 was completed based on the participant’s self-reported responses. For participants with both informant-rated and self-rated AD8 scores, the informant score was used. Participants were determined to be clinically impaired if they scored ≥2 on the informant-rated AD8 or ≥1 on the self-rated AD8 (maximum score of 8; higher scores indicate greater impairment) [41].

Eligible participants were scheduled for an in-person study visit at the Washington University Clinical and Translational Research Unit to complete cognitive screening (MoCA), a 60 mL blood collection, and a post-visit survey. The MoCA version 8.1 was administered by a trained research coordinator. A participant’s clinical cognitive status was determined by a composite of the MoCA and the AD8 that was obtained via the initial phone interview: an abnormal AD8 and MoCA of <26, or normal AD8 and MoCA of <24, was considered clinically impaired. A registered nurse or phlebotomist performed the blood collection per standard clinical protocol. Blood was collected in 10 mL EDTA tubes and centrifuged. Plasma was aliquoted and stored at -80°C until analysis via immunoprecipitation-mass spectrometry as previously described [13]. The buffy coat was sent to the Washington University Hope Center DNA and RNA Core for *APOE* genotyping. At the end of the visit, participants completed a computer survey about their study experience and opinions about the AD blood test. Survey questions about study experience were adapted from the Perceived Research Burden Assessment (PeRBA), a questionnaire measuring participants’ perceptions of burden in AD research [42].

The study was designed for approximately 25% of the sample of 1120 participants to be included in a confirmatory sub-study, which underwent a 400 mL blood collection for reproducibility, amyloid PET and magnetic resonance imaging (MRI) for validation of the blood test, and Clinical Dementia Rating® (CDR) for validation of the cognitive screening measures [43]. To efficiently power statistical analyses, the confirmatory sub-study was enriched for individuals with a positive AD blood test, cognitive impairment, and *APOE* ε4 positivity. Participants who were selected for the confirmatory sub-study were screened over the phone for contraindications to amyloid PET, brain MRI, and collection of 400 mL of blood. Participants without contraindications were invited for an in-person confirmatory visit. Participants with contraindications to MRI only were invited for an in-person confirmatory visit without the MRI component.

The time between a participant’s initial and confirmatory visits was planned for three months, but due to limited in-person visits related to the COVID-19 pandemic, this time interval was not restricted.

The 400 mL blood collection was performed at the Washington University Clinical and Translational Research Unit by a nurse or phlebotomist per standard clinical protocol. Participants completed a 30-minute amyloid PET scan using [11C] PiB (Pittsburgh Compound-B) and structural MRI scan at 3 Tesla (3T) on a combined PET-MRI scanner (Siemens Biograph mMR) at the Washington University Center for Clinical Imaging and Research. For those participants unable to undergo MRI due to conditions such as pacemaker or severe claustrophobia, PET-CT (Siemens Biograph mCT) was performed as a standalone exam without MRI. Participants in the confirmatory group were also invited to complete a phone-based CDR. The CDR was performed by seven different raters who completed online training and certification and held clinical coordinator, nurse coordinator, research assistant, or medical student roles.

### 2.3. Statistical Analyses

Differences between the characteristics of enrolled and expected groups were evaluated with standardized differences. The threshold for evaluation of differences used was 0.1 [44]. The area under the receiver operating characteristic curve (AUC) was used to evaluate the accuracy of the AD8 and MoCA in classifying CDR=0 and CDR>0 groups. Optimal cutoffs for the AD8 and MoCA were defined by the maximum combined sensitivity and sensitivity (Youden Index). Ordinal logistic regression was used to evaluate differences in survey responses as a function of age, self-identified race, educational background, and clinical status. Statistical analyses were performed using SAS version 9.4, and plots were created using R version 4.0.5. A *p* value of <0.05 was considered statistically significant.

## 3. Results

### 3.1. Participant characteristics and enrollment

Of the 1,699 individuals who were contacted about the study during the study period, 1,106 were screened (**Figure 1**). Only 52 individuals (4.7%) were ineligible for the study and 44 (4.0%) refused participation. An additional 151 individuals (13.7%) could not be re-contacted after expressing initial interest, had scheduling issues, or are being scheduled at the time of submission. The majority of individuals screened agreed to participate, and 859 (77.7%) were enrolled in SEABIRD and completed the initial visit between April 9, 2019 and October 31, 2022. Of 268 participants screened for the confirmatory sub-study, 11 (4.1%) were ineligible, 39 (14.6%) refused participation, and 44 (16.4%) were lost to follow-up or had scheduling issues (some will be scheduled for future visits). Notably, 22 of the 39 individuals (56.4%) who refused participation cited concerns about imaging (unwilling to be imaged or self-reported claustrophobia). A total of 149 individuals completed both amyloid PET and collection of 400 mL of blood. Some individuals underwent amyloid PET but have yet to complete blood collection, or vice versa. The time between participants’ initial visit and amyloid PET scan ranged from 39 days to 931 days (median 261 days) due to COVID-19 restrictions and scheduling limitations related to PET scan availability.

**Figure 1.**
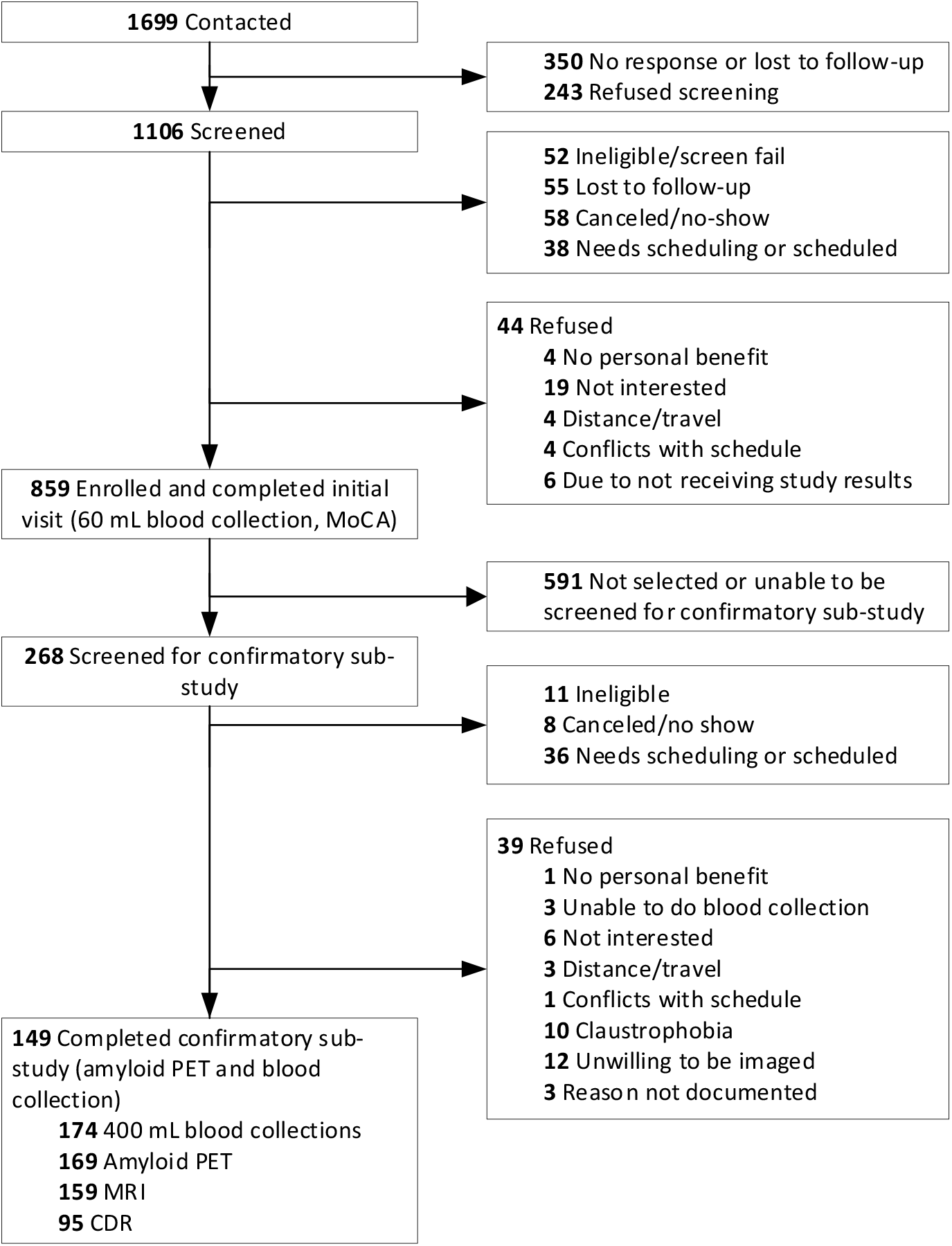
SEABIRD study flow. Abbreviations: MoCA, Montreal Cognitive Assessment; PET, positron emission tomography; MRI, magnetic resonance imaging; CDR, Clinical Dementia Rating.

Compared to the Saint Louis metropolitan area population, enrolled participants were somewhat more likely to be female and aged 70-79 years (**Table 1** and **Figure 2**). The percentage of *APOE* ε4 carriers was as expected for a general population, while cognitive status was more likely to be abnormal. The percentage of enrolled participants who identified their race as Black or African American was 20.6%, greater than expected (14.2%). Notably, SEABIRD participants were more highly educated than expected: they were less likely to have a high school degree or less (-0.50 standard difference), and much more likely to have a postgraduate degree (0.70 standard difference). Additionally, SEABIRD participants were less likely to report high cholesterol, diabetes, heart attack, kidney disease, and stroke, and more likely to report depression compared with the population of Missouri adults aged 65 and older.

**Table 1.**
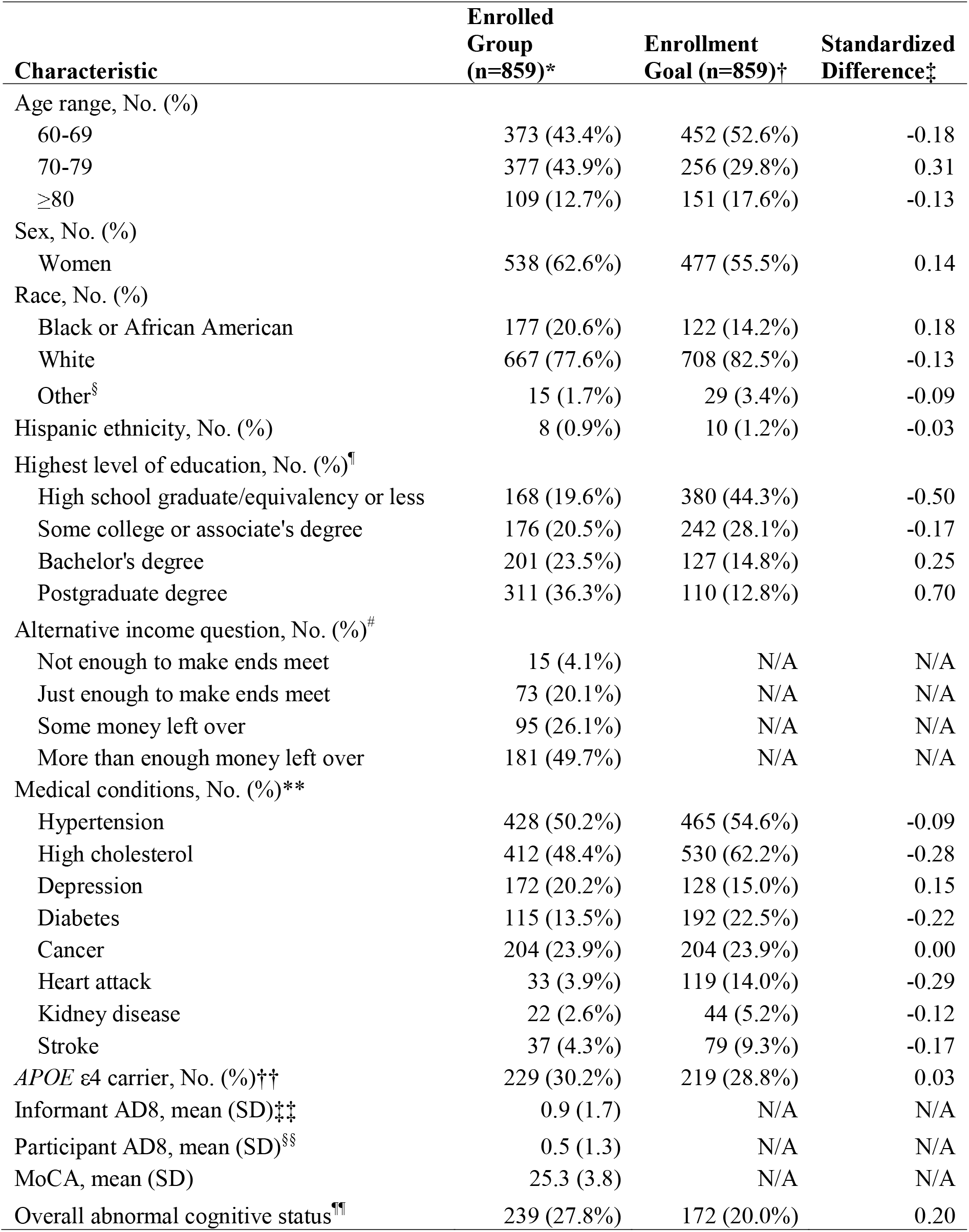
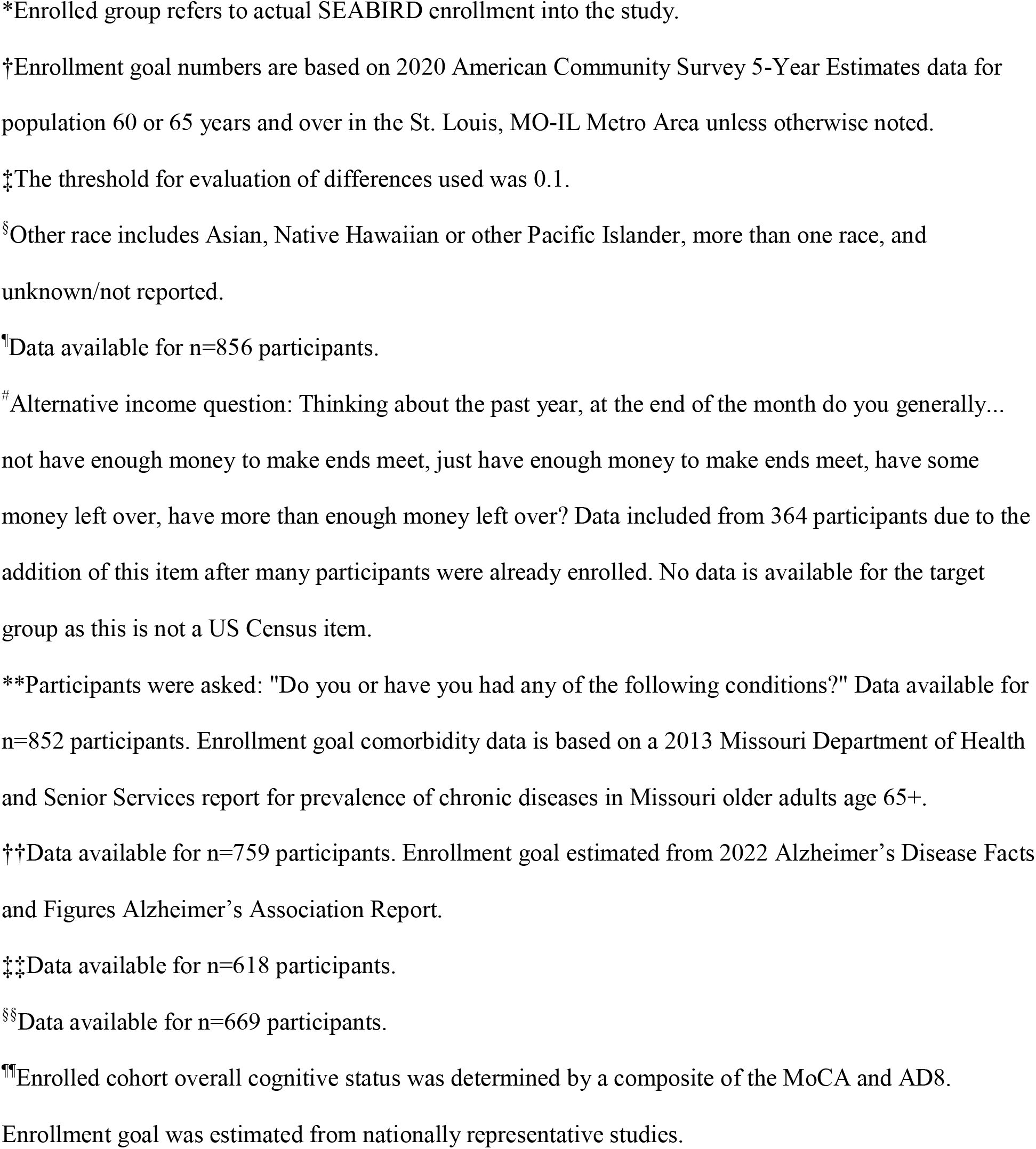
SEABIRD participant characteristics.

| **Characteristic** | **Enrolled Group (n=859)\*** | **Enrollment Goal (n=859)†** | **Standardized Difference‡** |
| --- | --- | --- | --- |
| Age range, No. (%) |  |  |  |
| 60-69 | 373 (43.4%) | 452 (52.6%) | -0.18 |
| 70-79 | 377 (43.9%) | 256 (29.8%) | 0.31 |
| ≥80 | 109 (12.7%) | 151 (17.6%) | -0.13 |
| Sex, No. (%) |  |  |  |
| Women | 538 (62.6%) | 477 (55.5%) | 0.14 |
| Race, No. (%) |  |  |  |
| Black or African American | 177 (20.6%) | 122 (14.2%) | 0.18 |
| White | 667 (77.6%) | 708 (82.5%) | -0.13 |
| Other§ | 15 (1.7%) | 29 (3.4%) | -0.09 |
| Hispanic ethnicity, No. (%) | 8 (0.9%) | 10 (1.2%) | -0.03 |
| Highest level of education, No. (%)¶ |  |  |  |
| High school graduate/equivalency or less | 168 (19.6%) | 380 (44.3%) | -0.50 |
| Some college or associate's degree | 176 (20.5%) | 242 (28.1%) | -0.17 |
| Bachelor's degree | 201 (23.5%) | 127 (14.8%) | 0.25 |
| Postgraduate degree | 311 (36.3%) | 110 (12.8%) | 0.70 |
| Alternative income question, No. (%)# |  |  |  |
| Not enough to make ends meet | 15 (4.1%) | N/A | N/A |
| Just enough to make ends meet | 73 (20.1%) | N/A | N/A |
| Some money left over | 95 (26.1%) | N/A | N/A |
| More than enough money left over | 181 (49.7%) | N/A | N/A |
| Medical conditions, No. (%)** |  |  |  |
| Hypertension | 428 (50.2%) | 465 (54.6%) | -0.09 |
| High cholesterol | 412 (48.4%) | 530 (62.2%) | -0.28 |
| Depression | 172 (20.2%) | 128 (15.0%) | 0.15 |
| Diabetes | 115 (13.5%) | 192 (22.5%) | -0.22 |
| Cancer | 204 (23.9%) | 204 (23.9%) | 0.00 |
| Heart attack | 33 (3.9%) | 119 (14.0%) | -0.29 |
| Kidney disease | 22 (2.6%) | 44 (5.2%) | -0.12 |
| Stroke | 37 (4.3%) | 79 (9.3%) | -0.17 |
| *APOE* ε4 carrier, No. (%)†† | 229 (30.2%) | 219 (28.8%) | 0.03 |
| Informant AD8, mean (SD)‡‡ | 0.9 (1.7) | N/A | N/A |
| Participant AD8, mean (SD)§§ | 0.5 (1.3) | N/A | N/A |
| MoCA, mean (SD) | 25.3 (3.8) | N/A | N/A |
| Overall abnormal cognitive status¶¶ | 239 (27.8%) | 172 (20.0%) | 0.20 |

\*Enrolled group refers to actual SEABIRD enrollment into the study.
†Enrollment goal numbers are based on 2020 American Community Survey 5-Year Estimates data for population 60 or 65 years and over in the St. Louis, MO-IL Metro Area unless otherwise noted.
‡The threshold for evaluation of differences used was 0.1.
§Other race includes Asian, Native Hawaiian or other Pacific Islander, more than one race, and unknown/not reported.
¶Data available for n=856 participants.

**Figure 2.**
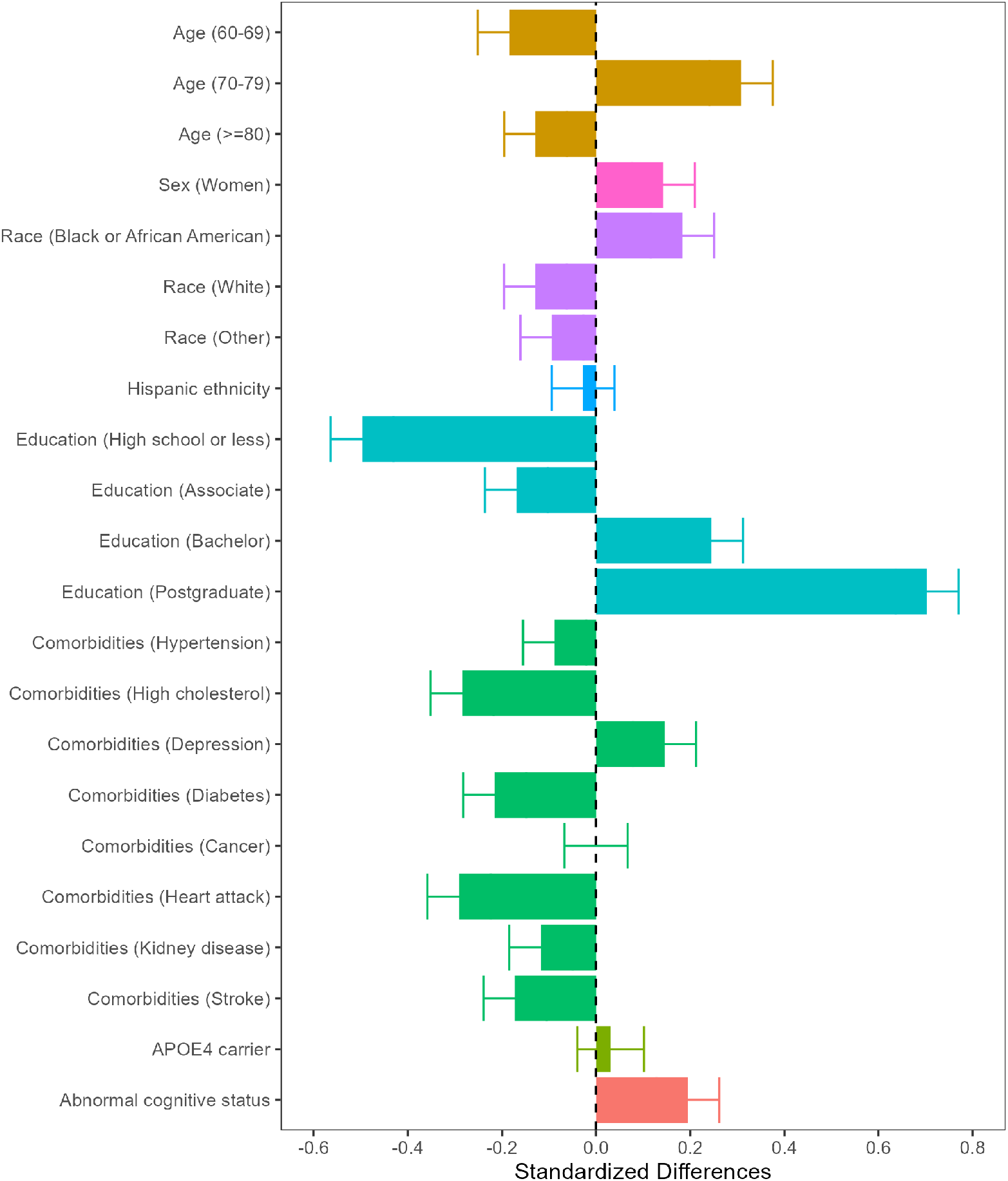
Characteristics of enrolled participants compared with enrollment goals. The SEABIRD enrollment characteristics compared to the local population are shown for age, sex, race, ethnicity, education, medical conditions, genetic APOE ε4 carrier status, and cognitive status. The SEABIRD study population is similar to the local population with respect to Hispanic ethnicity, APOE4 carrier status, and some comorbidities such as cancer. However, the study population has fewer than expected individuals with lower education, and other comorbidities such as stroke, kidney disease, and diabetes. In contrast, the study population includes greater than expected individuals with Black or African American race, eighth decade age group, depression, and cognitive impairment.

**Figure 3A** shows the number of participants enrolled from each recruitment source, and **Figure 3B-D** shows heat maps of the distribution of enrolled participants from different racial, age, and educational attainment groups. Participant registries and referrals from collaborating studies yielded participants who typically were White, older than 80 years, and highly educated. Word of mouth yielded participants who typically were non-White, older than 80 years, and less educated. Participants recruited through selective EMR review or direct physician referral typically were non-White, under age 70 years, and less educated. An article in the St. Louis Post Dispatch newspaper resulted in much lower recruitment of Black participants than advertisements in the St. Louis American, a newspaper with a wide African American readership.

**Figure 3.**
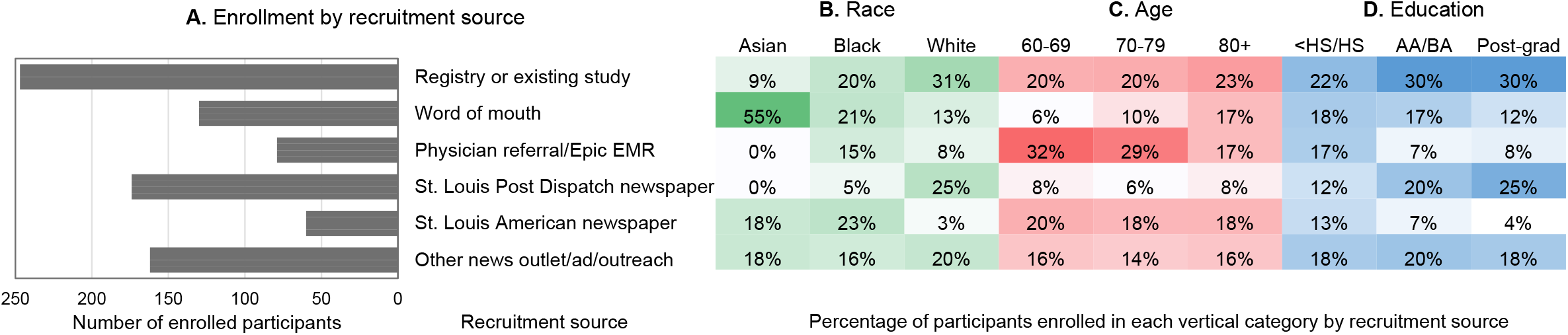
SEABIRD enrollment by recruitment sources. Recruitment sources and frequency of enrolled participants from each source are described in **(A)**. The distribution of participants from racial **(B)**, age **(C)**, and educational attainment **(D)** groups from each recruitment source are shown in heat maps. These findings indicate differential relative recruitment sources by race, age, and education. For example, word of mouth was higher in the 80+ compared to 60s age group, while physician referral was higher in the 60s and 70s compared to the 80+ age group. The St. Louis Post Dispatch had higher post-graduate education compared to high school education, and the St. Louis American newspaper had higher Asian and Black compared to White race enrollment. Abbreviations: EMR, electronic medical records; HS, high school; AA/BA, associate or bachelor of arts (some college, technical school, or college degree).

### 3.2. Clinical and cognitive results

Ninety-five participants in the confirmatory group completed a CDR assessment. The AUC for distinguishing cognitively unimpaired (CDR=0) from cognitively impaired (CDR>0) individuals was 0.72 for the informant-rated AD8, 0.46 for the self-rated AD8, 0.70 for the MoCA, and 0.79 for the AD8/MoCA composite (**Figure 4A**). The sensitivity was highest for the MoCA (0.83), while the informant-rated AD8 had the highest specificity (0.78). The optimal cutoff for the MoCA was the same as the standard assessment cutoff, while the optimal cutoff for the informant AD8 was one point lower than the standard cutoff of ≥2. An optimal cutoff was not generated for the self-rated AD8 as the AUC was less than 0.5. The probability of impairment on the AD8 and MoCA by cognitive status (CDR=0 or >0) is shown in **Figure 4B-C**. There was significant overlap between CDR groups in both the AD8 and MoCA scores.

**Figure 4.**
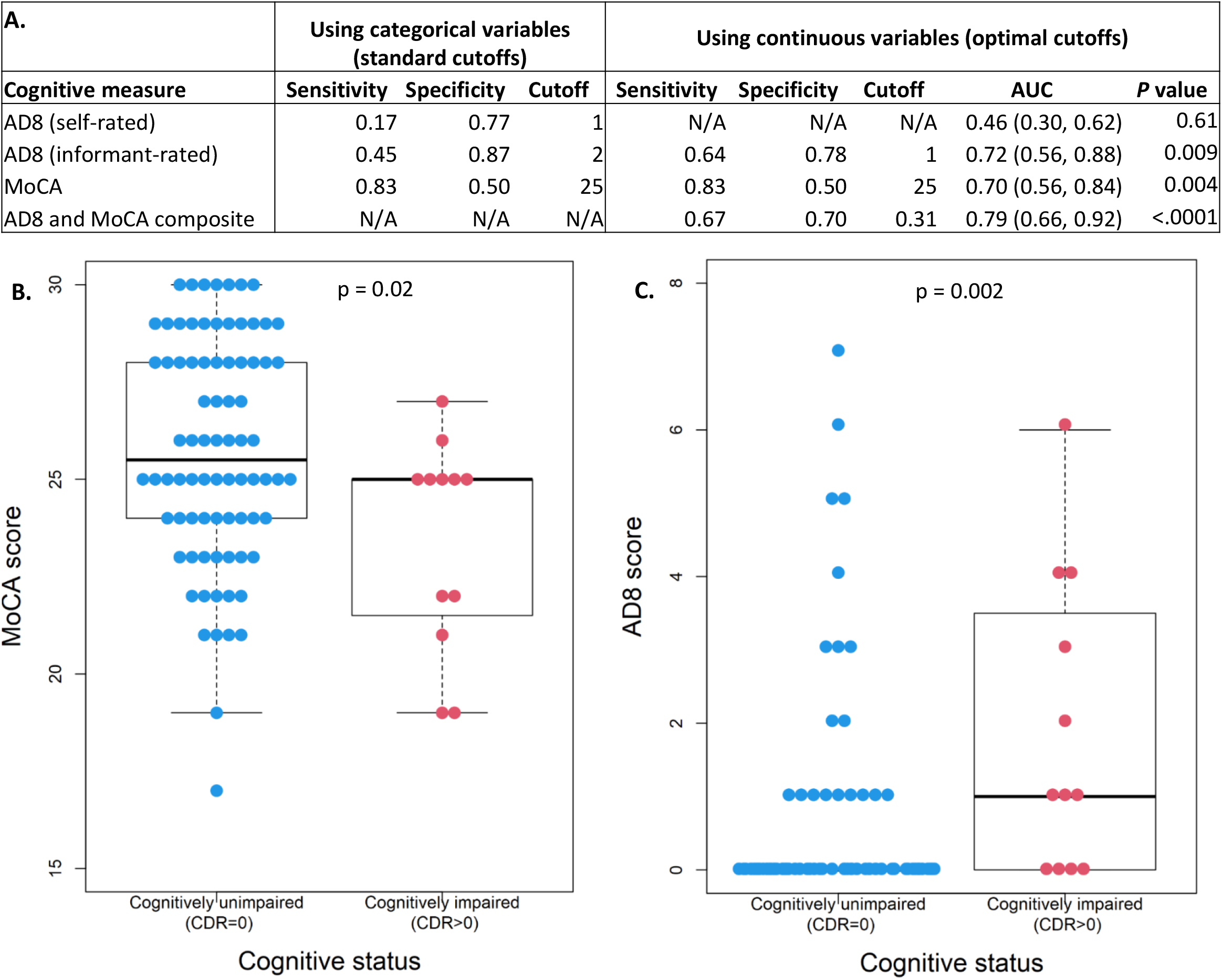
Concordance of AD8 and MoCA with CDR. N=95 participants had CDR scores. The informant AD8 and MoCA each have relatively modest accuracy of AUC of ≈0.7, while combining both measures in a composite modestly improved AUC to 0.79 **(A)**. Significant overlap between CDR groups was found for both the AD8 and MoCA **(B and C)**. The AD8 and MoCA composite in **(A)** was generated by first calculating the mean and SD of the CDR group and using these values to standardize the AD8 and MoCA. The MoCA score was inversed to be consistent with the AD8 (higher value means worse cognitive performance). AD8 and MoCA composite 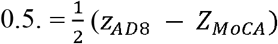. A *p* value of <0.05 indicates the AUC is significantly different from 0.5. For **(C)**, the informant AD8 was used when available, and the participant AD8 was used if the informant AD8 was missing. For the CDR=0 group, the box is not shown because the median, 25^th^, and 75^th^ quartile were all zero. Abbreviations: MoCA, Montreal Cognitive Assessment; AUC, area under the curve.

### 3.3. Participant perceptions of SEABIRD and the AD blood test

Figure 5. shows the results of the participant survey from the initial study visit. Overall, SEABIRD participants reported positive study experiences: most participants strongly disagreed that the study visit took too much time, or that study procedures caused distress. Generally, participants strongly agreed with statements that they were fairly compensated for their participation, able to get to the research site without difficulty, and had their questions and concerns addressed. Most participants also expressed willingness to consider participation in future AD studies. Participants generally expressed positive attitudes toward the AD blood test: they were willing to undergo an AD blood test for clinical trial screening, for clinical diagnosis (if symptomatic), or for risk stratification (if asymptomatic). Most participants strongly agreed that they would be interested in receiving their blood test results in the future. The frequency of responses for each survey item is shown in **Supplementary Table S1**.

**Figure 5.**
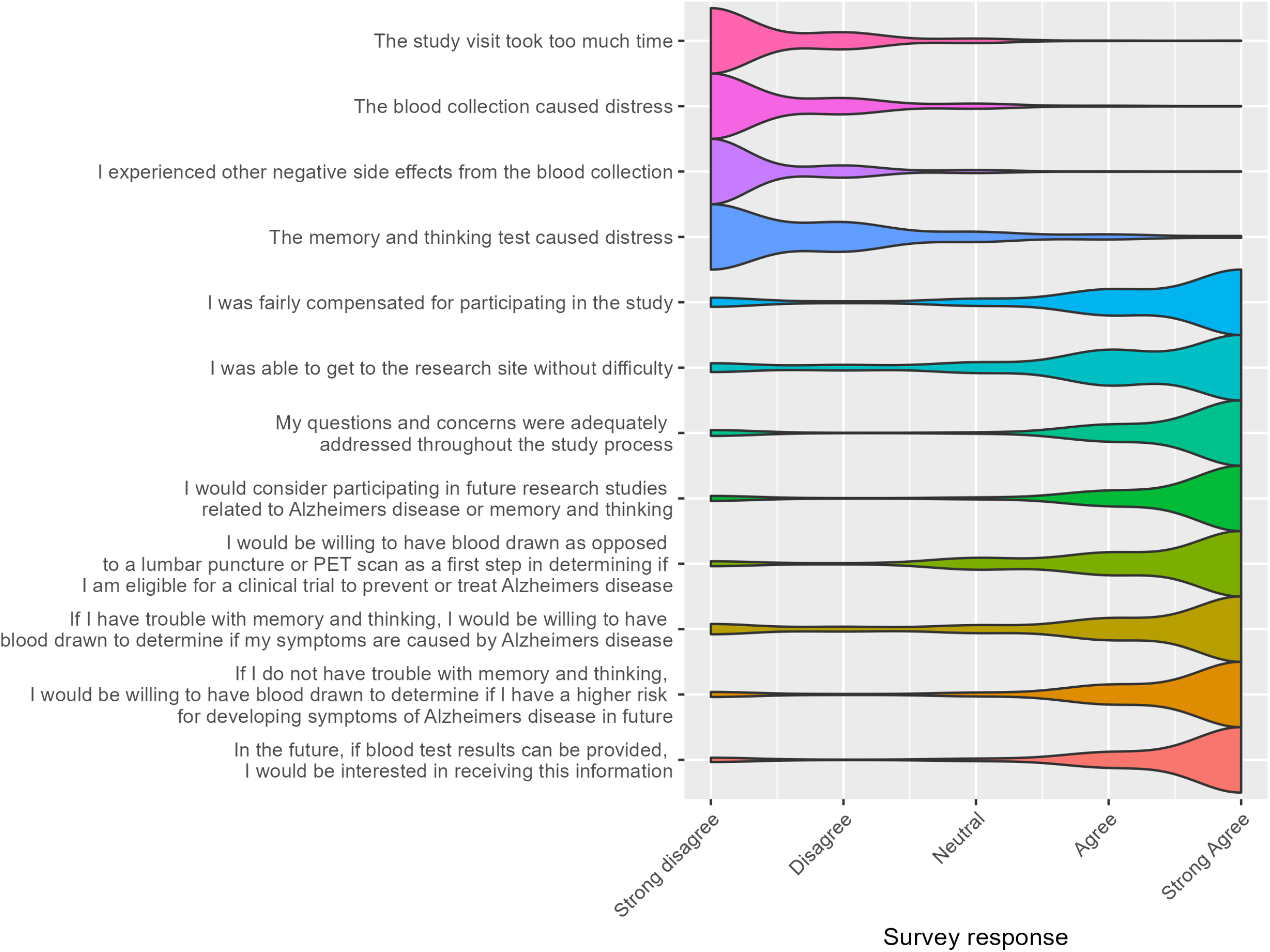
Participant survey results for initial study visit. Responses were on a 5-point Likert scale ranging from 1 (strongly disagree) to 5 (strongly agree). SEABIRD participants generally reported a positive study experience and positive attitudes toward the AD blood test.

Factors that influenced the experiences of participants were evaluated. Participants who agreed that the study visit took too much time or caused distress were more likely to be Black (compared to White), have a high school education or less (compared to a bachelor’s degree or higher), or have cognitive impairment (compared to cognitively unimpaired) (**Supplementary Table S2**). Participants who agreed that they were fairly compensated, able to get to the research site without difficulty, had their questions adequately addressed, or would consider participating in future AD studies were more likely to be age 60-69 years (compared to over age 80 years), White (compared to Black), or have a bachelor’s degree or higher (compared to a high school education or less). There were no significant differences in perceived study burden between participants with and without medical conditions (**Supplementary Table S4**). White, highly educated, younger, and cognitively unimpaired individuals tended to have more positive perceptions of the AD blood test: they were more likely to consider having blood drawn for clinical trial screening, having the blood test when asymptomatic to determine AD risk, and receiving results of the blood test in the future (**Supplementary Table S2**).

## 4. Discussion

In this ongoing observational study to validate an AD blood test, we demonstrated the feasibility of enrolling a diverse, community-based sample. The majority of screened participants enrolled in the study. The AD blood test and other study procedures were well accepted. Results from this study may provide insight into designing future studies to evaluate AD blood biomarkers in population-based studies, implement blood screening tests in clinical trials, and offer diagnostic blood tests in clinical settings.

A variety of recruitment avenues were used, and continued monitoring of recruitment demographics from different sources facilitated meeting study goals. The study was successful in recruiting a racially diverse sample compared to many AD studies [17], but was less successful in enrolling individuals with lower educational levels. One consequence of the COVID-19 pandemic was the nearly total cessation of in-person outreach. Consequently, the majority of recruitment focused on reaching potential participants electronically or through print, such as emails, flyers, and newspaper ads. These recruitment avenues could have led to over-representation of participants with higher education and literacy levels.

Enrollment of participants with some medical conditions, such as diabetes, heart attack, kidney disease, and stroke, was lower than expected. Participants with greater disease burden may experience greater difficulties participating in research and traveling to research centers, although our analysis did not show a significant difference in perceived study burden between participants with and without medical conditions. However, all data were collected from individuals who agreed to participate and therefore may not be representative of participants with medical conditions who did not agree to screening or enrollment.

The cognitive composite measure including the AD8 and MoCA was formulated to incorporate both a subjective and objective assessment, with the intent of minimizing over-reporting of clinical symptoms by the “worried well” and under-reporting by those who lack insight into cognitive impairment. Participants with normal AD8 scores and MoCA scores of <24 were considered cognitively impaired; this lower cutoff was selected for the MoCA because studies show the MoCA may over-classify participants as abnormal in community settings [45–47]. To maximize inclusion of underrepresented groups in the study, participants were not required to have a study partner. Therefore, the self-rated AD8 was used when an informant AD8 was unavailable although its concordance with CDR is lower than the informant-rated AD8 [41]. To our knowledge, this was the first time the self-rated AD8 was used in a community research setting, over the phone, in a diverse population. The concordance of the self-rated AD8 with CDR status was very poor (AUC 0.46). The informant-rated AD8 performed better when using an optimal cutoff of ≥1 to denote impairment rather than the standard cutoff of ≥2, possibly suggesting that a lower cutoff has more utility in a diverse community setting. The standard MoCA cutoff of <26 agreed with the optimal cutoff found in this cohort.

These results indicate the continued need to establish clinical and cognitive assessments that are rapid, minimally burdensome, valid, and precise. Misdiagnoses are common even with a comprehensive expert assessment: in a study evaluating the clinical diagnostic accuracy of AD dementia compared with different neuropathological criteria at 30 National Institute on Aging Alzheimer’s Disease Centers (ADCs), sensitivity ranged from 71% to 87% and specificity ranged from 44% to 71% [48]. Clinical diagnosis is aided by biological measures of AD pathology in populations with low comorbid burden that are typical of ADC cohorts. When participants have medical conditions that may impact cognition, biomarker testing is even more important in making an accurate etiological diagnosis.

Participant survey results demonstrated that use of AD blood tests is feasible and well-accepted in a diverse population. In fact, more participants reported distress caused by the cognitive assessments than by the blood collection. Although SEABIRD participants generally tolerated study procedures well and did not report excessive burden related to the study, Black individuals and those with lower education were more likely to perceive the study as burdensome, consistent with previous research [16]. Older and cognitively impaired participants were also more likely to experience study-associated burden and less likely to feel that they were fairly compensated for the study as has been reported in other studies [49]. Successful recruitment and retention of underrepresented groups requires continual efforts to understand barriers to research participation and incorporate feedback into study design.

The vast majority of SEABIRD participants were open to undergoing the AD blood test in research and clinical settings and to receiving study results, although there were some differences by race, educational level, and cognitive status. Black participants were less willing than White participants to undergo an AD blood test if symptomatic for purposes of clinical diagnosis, although there were no significant differences in responses by age, education, and cognitive status. To maximize the benefits of AD blood tests for all individuals, more research is needed to understand why Black participants may be less willing to undergo these tests.

There were multiple limitations in these analyses. Data on medical conditions were obtained through participant self-report, which may vary greatly from medical record data, especially for participants with cognitive impairment [50]. Acquisition of medical history data from participants’ medical records is planned to explore the relationship between specific medical conditions and AD biomarkers with greater accuracy. To minimize study burden, SEABIRD participants did not undergo a comprehensive clinical assessment by a dementia specialist. The accuracy of the CDR as a reference standard may be limited, and only 95 participants have completed the CDR to date.

In conclusion, SEABIRD demonstrates that it is feasible to study an AD blood test in a diverse population. The efficiency and scalability of the blood test enabled a single site to enroll 859 participants in 3.5 years, despite the occurrence of a global pandemic during much of the study period. The absence of a comprehensive clinical assessment, rather than being a limitation, may be a step toward a screening paradigm that prioritizes disease pathology over clinical symptomatology to accurately identify individuals who would be most likely to benefit from disease-modifying treatments. After enrollment and study visits are completed, the blood biomarker and imaging data from SEABIRD will be analyzed to further explore the relationships between cognition, amyloid pathology, and individual level factors such as age, race, *APOE* genotype, education, and medical conditions.

## Data Availability

All data produced in the present study are available upon reasonable request to the authors.

## Acknowledgments

We thank the participants and their families for their contributions to this study. We thank Melinda Hamilton, Melanie Burton, Diane Salamon, Melissa Sullivan, Tinishia Greene, and Derica Cartwright for their efforts in recruitment and study coordination; LaKisha Lloyd and Sheetal Mishall for coordinating imaging visits; Cynthia Hodge for administrative management; Paige Lawler and Lisa Soke for assistance with data quality control; and the nurses and medical assistants at the Washington University Clinical Translational Research Unit and Center for Clinical Imaging Research for their assistance during the study visits.

