## Supplemental tables for "Design and feasibility of an Alzheimer’s disease blood test study in a diverse community-based population"

**Table S1. Participant survey results by frequency of response**

| Survey question | Survey responses, frequency (%) | | | | | |
|---|---|---|---|---|---|---|
| | Strongly disagree | Disagree | Neutral | Agree | Strongly agree | Missing |
| The study visit took too much time. | 636 (74.56) | 164 (19.23) | 43 (5.04) | 5 (0.59) | 5 (0.59) | 6 |
| The blood collection caused distress. | 632 (74.62) | 154 (18.18) | 49 (5.79) | 9 (1.06) | 3 (0.35) | 12 |
| I experienced other negative side effects from the blood collection. | 676 (80.19) | 126 (14.95) | 32 (3.8) | 5 (0.59) | 4 (0.47) | 16 |
| The memory and thinking test caused distress. | 502 (58.99) | 221 (25.97) | 76 (8.93) | 38 (4.47) | 14 (1.65) | 8 |
| I was fairly compensated for participating in the study. | 71 (8.36) | 13 (1.53) | 55 (6.48) | 199 (23.44) | 511 (60.19) | 10 |
| I was able to get to the research site without difficulty. | 60 (7.03) | 37 (4.34) | 71 (8.32) | 239 (28.02) | 446 (52.29) | 6 |
| My questions and concerns were adequately addressed throughout the study process. | 56 (6.57) | 3 (0.35) | 12 (1.41) | 157 (18.43) | 624 (73.24) | 7 |
| I would consider participating in future research studies related to Alzheimer's disease or memory and thinking. | 48 (5.63) | 5 (0.59) | 18 (2.11) | 146 (17) | 637 (74.68) | 6 |
| I would be willing to have blood drawn as opposed to a lumbar puncture or PET scan as a first step in determining if I am eligible for a clinical trial to prevent or treat Alzheimer's disease. | 40 (4.7) | 9 (1.06) | 94 (11.05) | 178 (20.92) | 530 (62.28) | 8 |
| If I have trouble with memory and thinking, I would be willing to have blood drawn to determine if my symptoms are caused by Alzheimer's disease. | 80 (9.43) | 36 (4.25) | 60 (7.08) | 165 (19.46) | 507 (59.79) | 11 |
| If I do not have trouble with memory and thinking, I would be willing to have blood drawn to determine if I have a higher risk for developing symptoms of Alzheimer's disease in the future. | 45 (5.33) | 8 (0.95) | 33 (3.91) | 172 (20.36) | 587 (69.47) | 14 |
| In the future, if blood test results can be provided, I would be interested in receiving this information. | 41 (4.88) | 4 (0.48) | 23 (2.73) | 145 (17.24) | 628 (74.67) | 18 |

**Table S2. Survey results by age group, race, education, and cognitive status**

| Survey question | Test Group | Reference Group | Odds ratio | Lower CI | Upper CI | p value | |
|---|---|---|---|---|---|---|---|
| 1. The study visit took too much time. | Age 70-79 | Age 60-69 | 0.994 | 0.713 | 1.387 | 0.9731 | * |
| | Age 80+ | Age 60-69 | 1.561 | 0.987 | 2.467 | 0.0568 | |
| | Other race | Black | 2.168 | 0.792 | 5.937 | 0.1322 | |
| | White | Black | **0.627** | **0.437** | **0.899** | **0.0110** | |
| | Some college/associate degree | High school or less | 0.686 | 0.434 | 1.083 | 0.1060 | * |
| | Bachelor's degree | High school or less | **0.424** | **0.264** | **0.682** | **0.0004** | |
| | Postgraduate degree | High school or less | **0.615** | **0.41** | **0.922** | **0.0188** | |
| | Cognitively impaired | Cognitively unimpaired | **2.087** | **1.51** | **2.884** | **<.0001** | |
| 2. The blood collection caused distress. | Age 70-79 | Age 60-69 | 0.884 | 0.637 | 1.227 | 0.4620 | |
| | Age 80+ | Age 60-69 | 0.841 | 0.511 | 1.384 | 0.4960 | |
| | Other race | Black | 0.68 | 0.209 | 2.208 | 0.5206 | |
| | White | Black | **0.566** | **0.397** | **0.808** | **0.0017** | |
| | Some college/associate degree | High school or less | 0.545 | 0.341 | 0.873 | 0.0116 | * |
| | Bachelor's degree | High school or less | **0.405** | **0.251** | **0.653** | **0.0002** | |
| | Postgraduate degree | High school or less | **0.669** | **0.447** | **1** | **0.0499** | |
| | Cognitively impaired | Cognitively unimpaired | **1.501** | **1.079** | **2.089** | **0.0159** | |
| 3. I experienced other negative side effects from the blood collection. | Age 70-79 | Age 60-69 | 0.941 | 0.656 | 1.35 | 0.7414 | |
| | Age 80+ | Age 60-69 | 0.966 | 0.563 | 1.658 | 0.9006 | |
| | Other race | Black | 1.128 | 0.371 | 3.428 | 0.8321 | |
| | White | Black | **0.474** | **0.324** | **0.693** | **0.0001** | |
| | Some college/associate degree | High school or less | 0.565 | 0.341 | 0.935 | 0.0264 | * |
| | Bachelor's degree | High school or less | **0.406** | **0.242** | **0.68** | **0.0006** | |
| | Postgraduate degree | High school or less | **0.568** | **0.366** | **0.88** | **0.0113** | |
| | Cognitively impaired | Cognitively unimpaired | **1.964** | **1.379** | **2.797** | **0.0002** | |
| 4. The memory and thinking test caused distress. | Age 70-79 | Age 60-69 | 0.898 | 0.677 | 1.19 | 0.4526 | |
| | Age 80+ | Age 60-69 | 0.889 | 0.581 | 1.361 | 0.5874 | |
| | Other race | Black | 1.143 | 0.421 | 3.108 | 0.7930 | |
| | White | Black | 0.803 | 0.581 | 1.109 | 0.1828 | |
| | Some college/associate degree | High school or less | 0.778 | 0.513 | 1.178 | 0.2354 | |
| | Bachelor's degree | High school or less | 0.807 | 0.541 | 1.204 | 0.2932 | |
| | Postgraduate degree | High school or less | 0.799 | 0.554 | 1.153 | 0.2300 | |
| | Cognitively impaired | Cognitively unimpaired | **2.031** | **1.521** | **2.713** | **<.0001** | |
| 5. I was fairly compensated for participating in the study. | Age 70-79 | Age 60-69 | 1.021 | 0.767 | 1.36 | 0.8860 | |
| | Age 80+ | Age 60-69 | **0.631** | **0.419** | **0.952** | **0.0282** | |
| | Other race | Black | 1.903 | 0.654 | 5.538 | 0.2378 | |
| | White | Black | **2.363** | **1.722** | **3.244** | **<.0001** | |
| | Some college/associate degree | High school or less | 1.036 | 0.687 | 1.562 | 0.8663 | * |
| | Bachelor's degree | High school or less | **1.608** | **1.065** | **2.426** | **0.0237** | |
| | Postgraduate degree | High school or less | 1.286 | 0.889 | 1.861 | 0.1821 | |
| | Cognitively impaired | Cognitively unimpaired | **0.421** | **0.315** | **0.564** | **<.0001** | |
| 6. I was able to get to the research site without difficulty. | Age 70-79 | Age 60-69 | 0.844 | 0.64 | 1.112 | 0.2285 | |
| | Age 80+ | Age 60-69 | **0.344** | **0.231** | **0.512** | **<.0001** | |
| | Other race | Black | 1.028 | 0.39 | 2.711 | 0.9559 | |
| | White | Black | **1.64** | **1.203** | **2.235** | **0.0018** | |
| | Some college/associate degree | High school or less | 0.985 | 0.661 | 1.468 | 0.9424 | |
| | Bachelor's degree | High school or less | 1.435 | 0.967 | 2.129 | 0.0726 | |
| | Postgraduate degree | High school or less | 1.07 | 0.75 | 1.527 | 0.7092 | |
| | Cognitively impaired | Cognitively unimpaired | **0.513** | **0.387** | **0.679** | **<.0001** | |
| 7. My questions and concerns were adequately addressed throughout the study process. | Age 70-79 | Age 60-69 | 1.025 | 0.739 | 1.422 | 0.8838 | |
| | Age 80+ | Age 60-69 | **0.564** | **0.361** | **0.881** | **0.0119** | |
| | Other race | Black | 1.297 | 0.45 | 3.735 | 0.6304 | * |
| | White | Black | **2.802** | **1.99** | **3.944** | **<.0001** | |
| | Some college/associate degree | High school or less | 1.214 | 0.784 | 1.881 | 0.3841 | * |
| | Bachelor's degree | High school or less | **2.467** | **1.548** | **3.93** | **0.0001** | |
| | Postgraduate degree | High school or less | **2.015** | **1.341** | **3.026** | **0.0007** | |
| | Cognitively impaired | Cognitively unimpaired | 0.339 | 0.247 | 0.465 | <.0001 | * |

| | | | | | | | |
|---|---|---|---|---|---|---|---|
| 8. I would consider participating in future research studies related to Alzheimer's disease or memory and thinking. | Age 70-79 | Age 60-69 | 0.929 | 0.664 | 1.299 | 0.6659 | * |
| | Age 80+ | Age 60-69 | **0.474** | **0.303** | **0.742** | **0.0011** | |
| | Other race | Black | 1.08 | 0.377 | 3.095 | 0.8858 | * |
| | White | Black | **2.547** | **1.799** | **3.607** | **<.0001** | |
| | Some college/associate degree | High school or less | 1.259 | 0.801 | 1.978 | 0.3183 | * |
| | Bachelor's degree | High school or less | **2.207** | **1.374** | **3.545** | **0.0011** | |
| | Postgraduate degree | High school or less | **1.705** | **1.13** | **2.573** | **0.0110** | |
| | Cognitively impaired | Cognitively unimpaired | **0.364** | **0.264** | **0.502** | **<.0001** | |
| 9. I would be willing to have blood drawn as opposed to a lumbar puncture or PET scan as a first step in determining if I am eligible for a clinical trial to prevent or treat Alzheimer's disease. | Age 70-79 | Age 60-69 | 1.259 | 0.94 | 1.686 | 0.1216 | |
| | Age 80+ | Age 60-69 | **0.652** | **0.433** | **0.982** | **0.0409** | |
| | Other race | Black | 1.641 | 0.568 | 4.741 | 0.3601 | * |
| | White | Black | **1.769** | **1.284** | **2.438** | **0.0005** | |
| | Some college/associate degree | High school or less | 1.023 | 0.678 | 1.543 | 0.9149 | * |
| | Bachelor's degree | High school or less | 1.436 | 0.954 | 2.161 | 0.0827 | |
| | Postgraduate degree | High school or less | **1.566** | **1.076** | **2.281** | **0.0193** | |
| | Cognitively impaired | Cognitively unimpaired | **0.633** | **0.472** | **0.848** | **0.0022** | |
| 10. If I have trouble with memory and thinking, I would be willing to have blood drawn to determine if my symptoms are caused by Alzheimer's disease. | Age 70-79 | Age 60-69 | 1.127 | 0.848 | 1.497 | 0.4096 | |
| | Age 80+ | Age 60-69 | 0.893 | 0.59 | 1.352 | 0.5942 | |
| | Other race | Black | 1.634 | 0.561 | 4.759 | 0.3682 | * |
| | White | Black | **1.427** | **1.036** | **1.964** | **0.0293** | |
| | Some college/associate degree | High school or less | 1.141 | 0.752 | 1.731 | 0.5339 | |
| | Bachelor's degree | High school or less | 1.157 | 0.772 | 1.733 | 0.4809 | |
| | Postgraduate degree | High school or less | 1.054 | 0.73 | 1.524 | 0.7777 | |
| | Cognitively impaired | Cognitively unimpaired | 0.859 | 0.641 | 1.151 | 0.3099 | |
| 11. If I do not have trouble with memory and thinking, I would be willing to have blood drawn to determine if I have a higher risk for developing symptoms of Alzheimer's disease in the future. | Age 70-79 | Age 60-69 | 1.114 | 0.814 | 1.525 | 0.4997 | * |
| | Age 80+ | Age 60-69 | **0.567** | **0.367** | **0.875** | **0.0104** | |
| | Other race | Black | 1.355 | 0.476 | 3.861 | 0.5692 | |
| | White | Black | **2.463** | **1.765** | **3.437** | **<.0001** | |
| | Some college/associate degree | High school or less | 1.278 | 0.834 | 1.959 | 0.2604 | |
| | Bachelor's degree | High school or less | **2.478** | **1.583** | **3.88** | **<.0001** | |
| | Postgraduate degree | High school or less | **1.856** | **1.257** | **2.74** | **0.0019** | |
| | Cognitively impaired | Cognitively unimpaired | **0.499** | **0.366** | **0.679** | **<.0001** | |
| 12. In the future, if blood test results can be provided, I would be interested in receiving this information. | Age 70-79 | Age 60-69 | 0.995 | 0.711 | 1.392 | 0.9767 | |
| | Age 80+ | Age 60-69 | **0.58** | **0.366** | **0.918** | **0.0200** | |
| | Other race | Black | 3.011 | 0.805 | 11.264 | 0.1016 | |
| | White | Black | **2.617** | **1.85** | **3.703** | **<.0001** | |
| | Some college/associate degree | High school or less | 1.379 | 0.882 | 2.157 | 0.1587 | * |
| | Bachelor's degree | High school or less | **3.064** | **1.884** | **4.985** | **<.0001** | |
| | Postgraduate degree | High school or less | **2.124** | **1.41** | **3.2** | **0.0003** | |
| | Cognitively impaired | Cognitively unimpaired | **0.501** | **0.362** | **0.694** | **<.0001** | |

*assumption not met

Ordinal logistic regressions were used to compare differences in survey responses. Higher OR indicates greater odds of rating a higher score (more agreement) for the Test Group vs. the Reference Group.

**Table S3. Survey results by individual medical condition**

| Survey question | Test Group | Reference Group | Odds ratio | Lower CI | Upper CI | p value | |
|---|---|---|---|---|---|---|---|
| 1. The study visit took too much time. | Hypertension | No hypertension | **1.360** | **1.000** | **1.849** | **0.0500** | |
| | High cholesterol | No high cholesterol | 1.125 | 0.829 | 1.528 | 0.4488 | |
| | Depression | No depression | 0.948 | 0.645 | 1.394 | 0.7868 | |
| | Diabetes | No diabetes | 1.248 | 0.811 | 1.920 | 0.3133 | * |
| | Cancer | No cancer | 1.272 | 0.897 | 1.803 | 0.1768 | |
| | Heart Attack | No heart attack | 1.849 | 0.895 | 3.819 | 0.0970 | |
| | Kidney disease | No kidney disease | 1.149 | 0.452 | 2.920 | 0.7712 | |
| | Stroke | No stroke | 1.068 | 0.505 | 2.258 | 0.8633 | |
| 2. The blood collection caused distress. | Hypertension | No hypertension | 1.181 | 0.869 | 1.606 | 0.2882 | |
| | High cholesterol | No high cholesterol | 0.978 | 0.720 | 1.330 | 0.8894 | |
| | Depression | No depression | 0.881 | 0.596 | 1.304 | 0.5273 | |
| | Diabetes | No diabetes | 0.913 | 0.579 | 1.441 | 0.6971 | * |
| | Cancer | No cancer | 0.860 | 0.594 | 1.244 | 0.4234 | |
| | Heart Attack | No heart attack | 1.833 | 0.887 | 3.786 | 0.1017 | |
| | Kidney disease | No kidney disease | 1.826 | 0.762 | 4.375 | 0.1768 | * |
| | Stroke | No stroke | 0.832 | 0.375 | 1.846 | 0.6514 | |
| 3. I experienced other negative side effects from the blood collection. | Hypertension | No hypertension | 1.196 | 0.853 | 1.677 | 0.2990 | |
| | High cholesterol | No high cholesterol | 0.909 | 0.648 | 1.274 | 0.5795 | |
| | Depression | No depression | 0.885 | 0.575 | 1.362 | 0.5792 | |
| | Diabetes | No diabetes | 0.950 | 0.578 | 1.562 | 0.8401 | * |
| | Cancer | No cancer | 0.916 | 0.612 | 1.370 | 0.6685 | |
| | Heart Attack | No heart attack | **2.415** | **1.155** | **5.046** | **0.0191** | |
| | Kidney disease | No kidney disease | 1.836 | 0.740 | 4.560 | 0.1902 | * |
| | Stroke | No stroke | 1.084 | 0.479 | 2.449 | 0.8470 | |
| 4. The memory and thinking test caused distress. | Hypertension | No hypertension | **1.313** | **1.007** | **1.711** | **0.0439** | |
| | High cholesterol | No high cholesterol | 0.963 | 0.739 | 1.254 | 0.7778 | |
| | Depression | No depression | 1.230 | 0.889 | 1.703 | 0.2117 | |
| | Diabetes | No diabetes | 0.954 | 0.647 | 1.407 | 0.8121 | |
| | Cancer | No cancer | 1.134 | 0.834 | 1.543 | 0.4227 | * |
| | Heart Attack | No heart attack | 1.819 | 0.935 | 3.539 | 0.0781 | |
| | Kidney disease | No kidney disease | **2.385** | **1.099** | **5.172** | **0.0278** | |
| | Stroke | No stroke | 0.820 | 0.417 | 1.612 | 0.5649 | |
| 5. I was fairly compensated for participating in the study. | Hypertension | No hypertension | **0.736** | **0.563** | **0.961** | **0.0241** | |
| | High cholesterol | No high cholesterol | 1.127 | 0.864 | 1.471 | 0.3781 | |
| | Depression | No depression | 0.949 | 0.682 | 1.322 | 0.7586 | |
| | Diabetes | No diabetes | 1.068 | 0.722 | 1.580 | 0.7410 | |
| | Cancer | No cancer | 1.348 | 0.978 | 1.859 | 0.0684 | |
| | Heart Attack | No heart attack | 1.722 | 0.786 | 3.773 | 0.1744 | |
| | Kidney disease | No kidney disease | 1.003 | 0.434 | 2.316 | 0.9944 | * |
| | Stroke | No stroke | 1.281 | 0.639 | 2.571 | 0.4852 | * |
| 6. I was able to get to the research site without difficulty. | Hypertension | No hypertension | 0.820 | 0.635 | 1.057 | 0.1260 | |
| | High cholesterol | No high cholesterol | 0.954 | 0.739 | 1.231 | 0.7170 | |
| | Depression | No depression | 0.839 | 0.612 | 1.149 | 0.2737 | |
| | Diabetes | No diabetes | 1.261 | 0.862 | 1.846 | 0.2321 | |
| | Cancer | No cancer | 1.064 | 0.788 | 1.436 | 0.6874 | |
| | Heart Attack | No heart attack | 0.650 | 0.336 | 1.255 | 0.1991 | |
| | Kidney disease | No kidney disease | 0.949 | 0.427 | 2.109 | 0.8975 | |
| | Stroke | No stroke | 0.826 | 0.443 | 1.540 | 0.5482 | |

| | | | | | | | |
|---|---|---|---|---|---|---|---|
| 7. My questions and concerns were adequately addressed throughout the study process. | Hypertension | No hypertension | 0.740 | 0.547 | 1.001 | 0.0505 | |
| | High cholesterol | No high cholesterol | 1.114 | 0.824 | 1.505 | 0.4831 | |
| | Depression | No depression | 1.230 | 0.834 | 1.814 | 0.2961 | |
| | Diabetes | No diabetes | 1.242 | 0.786 | 1.965 | 0.3534 | |
| | Cancer | No cancer | 1.252 | 0.869 | 1.804 | 0.2281 | |
| | Heart Attack | No heart attack | 0.946 | 0.429 | 2.088 | 0.8912 | |
| | Kidney disease | No kidney disease | 0.465 | 0.203 | 1.062 | 0.0691 | |
| | Stroke | No stroke | 1.040 | 0.485 | 2.230 | 0.9207 | |
| 8. I would consider participating in future research studies related to Alzheimer's disease or memory and thinking. | Hypertension | No hypertension | 0.746 | 0.549 | 1.015 | 0.0618 | |
| | High cholesterol | No high cholesterol | 1.303 | 0.957 | 1.773 | 0.0925 | |
| | Depression | No depression | **1.752** | **1.144** | **2.683** | **0.0099** | |
| | Diabetes | No diabetes | 1.342 | 0.833 | 2.161 | 0.2260 | * |
| | Cancer | No cancer | 1.129 | 0.783 | 1.627 | 0.5170 | |
| | Heart Attack | No heart attack | 0.990 | 0.438 | 2.236 | 0.9809 | |
| | Kidney disease | No kidney disease | **0.436** | **0.191** | **0.998** | **0.0493** | |
| | Stroke | No stroke | 0.878 | 0.420 | 1.835 | 0.7292 | * |
| 9. I would be willing to have blood drawn as opposed to a lumbar puncture or PET scan as a first step in determining if I am eligible for a clinical trial to prevent or treat Alzheimer's disease. | Hypertension | No hypertension | 0.858 | 0.656 | 1.124 | 0.2667 | |
| | High cholesterol | No high cholesterol | 1.098 | 0.838 | 1.437 | 0.4984 | |
| | Depression | No depression | 1.249 | 0.885 | 1.763 | 0.2052 | |
| | Diabetes | No diabetes | 0.967 | 0.654 | 1.431 | 0.8677 | * |
| | Cancer | No cancer | 1.050 | 0.765 | 1.443 | 0.7614 | |
| | Heart Attack | No heart attack | 1.520 | 0.698 | 3.310 | 0.2918 | |
| | Kidney disease | No kidney disease | 0.723 | 0.321 | 1.626 | 0.4323 | * |
| | Stroke | No stroke | 0.896 | 0.464 | 1.730 | 0.7438 | * |
| 10. If I have trouble with memory and thinking, I would be willing to have blood drawn to determine if my symptoms are caused by Alzheimer's disease. | Hypertension | No hypertension | 1.086 | 0.834 | 1.415 | 0.5395 | |
| | High cholesterol | No high cholesterol | 1.226 | 0.940 | 1.598 | 0.1326 | |
| | Depression | No depression | **1.771** | **1.241** | **2.528** | **0.0016** | |
| | Diabetes | No diabetes | 1.144 | 0.772 | 1.694 | 0.5024 | |
| | Cancer | No cancer | 1.076 | 0.787 | 1.470 | 0.6455 | |
| | Heart Attack | No heart attack | 1.950 | 0.874 | 4.351 | 0.1031 | |
| | Kidney disease | No kidney disease | 1.355 | 0.564 | 3.254 | 0.4971 | * |
| | Stroke | No stroke | 2.530 | 1.138 | 5.628 | 0.0228 | * |
| 11. If I do not have trouble with memory and thinking, I would be willing to have blood drawn to determine if I have a higher risk for developing symptoms of Alzheimer's disease in the future. | Hypertension | No hypertension | 0.823 | 0.617 | 1.099 | 0.1869 | |
| | High cholesterol | No high cholesterol | 1.136 | 0.850 | 1.516 | 0.3893 | |
| | Depression | No depression | **1.622** | **1.099** | **2.393** | **0.0148** | |
| | Diabetes | No diabetes | 1.019 | 0.668 | 1.553 | 0.9316 | |
| | Cancer | No cancer | 1.046 | 0.743 | 1.472 | 0.7967 | |
| | Heart Attack | No heart attack | 0.805 | 0.386 | 1.681 | 0.5642 | * |
| | Kidney disease | No kidney disease | 0.873 | 0.362 | 2.107 | 0.7625 | |
| | Stroke | No stroke | 1.928 | 0.830 | 4.477 | 0.1268 | * |
| 12. In the future, if blood test results can be provided, I would be interested in receiving this information. | Hypertension | No hypertension | 0.682 | 0.500 | 0.930 | 0.0156 | * |
| | High cholesterol | No high cholesterol | 1.058 | 0.777 | 1.441 | 0.7196 | |
| | Depression | No depression | 1.405 | 0.935 | 2.112 | 0.1018 | |
| | Diabetes | No diabetes | 1.186 | 0.743 | 1.892 | 0.4747 | |
| | Cancer | No cancer | 1.006 | 0.700 | 1.446 | 0.9732 | * |
| | Heart Attack | No heart attack | 1.051 | 0.460 | 2.403 | 0.9064 | |
| | Kidney disease | No kidney disease | 0.663 | 0.275 | 1.602 | 0.3618 | * |
| | Stroke | No stroke | 1.186 | 0.536 | 2.623 | 0.6744 | * |

*assumption not met
Ordinal logistic regressions were used to compare differences in survey responses. Higher OR indicates greater odds of rating a higher score (more agreement) for the Test Group vs. the Reference Group.

**Table S4. Survey results by presence or absence of medical conditions**

| Survey question | Test Group | Reference Group | Odds ratio | Lower CI | Upper CI | p value |
|---|---|---|---|---|---|---|
| The study visit took too much time. | One or more medical conditions | No medical conditions | 0.655 | 1.436 | 0.879 | 0.97 |
| The blood collection caused distress. | One or more medical conditions | No medical conditions | 0.621 | 1.356 | 0.6662 | 0.918 |
| I experienced other negative side effects from the blood collection. | One or more medical conditions | No medical conditions | 0.511 | 1.171 | 0.2246 | 0.773 |
| The memory and thinking test caused distress. | One or more medical conditions | No medical conditions | 0.991 | 2.008 | 0.056 | 1.411 |
| I was fairly compensated for participating in the study. | One or more medical conditions | No medical conditions | 0.723 | 1.431 | 0.9211 | 1.017 |
| I was able to get to the research site without difficulty. | One or more medical conditions | No medical conditions | 0.742 | 1.427 | 0.8642 | 1.029 |
| My questions and concerns were adequately addressed throughout the study process. | One or more medical conditions | No medical conditions | 0.846 | 1.79 | 0.2783 | 1.23 * |
| I would consider participating in future research studies related to Alzheimer's disease or memory and thinking. | One or more medical conditions | No medical conditions | 0.563 | 1.267 | 0.4143 | 0.845 * |
| I would be willing to have blood drawn as opposed to a lumbar puncture or PET scan as a first step in determining if I am eligible for a clinical trial to prevent or treat Alzheimer's disease. | One or more medical conditions | No medical conditions | 1.203 | 0.857 | 1.69 | 0.2857 |
| If I have trouble with memory and thinking, I would be willing to have blood drawn to determine if my symptoms are caused by Alzheimer's disease. | One or more medical conditions | No medical conditions | 0.335 | 0.958 | 1.861 | 0.0879 |
| If I do not have trouble with memory and thinking, I would be willing to have blood drawn to determine if I have a higher risk for developing symptoms of Alzheimer's disease in the future. | One or more medical conditions | No medical conditions | 1.26 | 0.88 | 1.805 | 0.2075 * |
| In the future, if blood test results can be provided, I would be interested in receiving this information. | One or more medical conditions | No medical conditions | 0.996 | 0.67 | 1.48 | 0.9845 * |

*assumption not met

Ordinal logistic regressions were used to compare differences in survey responses. Higher OR indicates greater odds of rating a higher score (more agreement) for the Test Group vs. the Reference Group.
